# GABA Probiotic Lactiplantibacillus plantarum Lp815 Improves Sleep, Anxiety and Increases Urinary GABA: a Randomized, Double-Blind, Placebo-Controlled Study

**DOI:** 10.1101/2025.04.14.25325830

**Authors:** Azure D. Grant, Marie Crisel B. Erfe, Camille J. Delebecque, David Keller, Noah P. Zimmerman, Amy Kazaryan, Paige L. Oliver, Jordan Moos, Veronica Luna, Noah Craft

**Author notes:** **Corresponding Author Contact** Noah Craft, MD, PhD h People Science, Inc. 3415 S. Sepulveda Blvd, Suite 1123, Los Angeles, CA 90034. Indicates co-first authorship (Azure D. Grant, Marie Crisel B. Erfe).

## Abstract

Gamma-aminobutyric acid, or GABA, the major inhibitory neurotransmitter, acts both through the central nervous system and via microbial production in the gut. Strains able to augment gut GABA production could be novel targets for modulation of sleep and mood. We previously demonstrated that *Lactiplantibacillus plantarum* Lp815 improves symptoms of anxiety. The current study investigates the impact of Lp815 on subjective sleep, mood, digestion and objective wearable sleep data over 6 weeks in adults with self-reported sleep disturbance. The trial was decentralized, double-blind, randomized and placebo controlled. Participants were screened for at least moderate symptoms of insomnia and blindly assigned to receive either a placebo capsule or 5 billion CFU of Lp815 per day. 139 Individuals (5 Billion CFU n=70, placebo n=69) were evaluated, aged 44 ± 13 years, 51.1% female and 55.4% Caucasian. A sub cohort (n=17, 5 Billion CFU n=9, Placebo n=8) submitted a urinary time series for neurotransmitter analysis. Participants who received 5 billion CFU exhibited significantly lower insomnia severity index scores at 6 weeks compared to placebo (p<0.05). This result was clinically meaningful, with 77.3% of participants in the 5 billion CFU cohort exhibiting improvement by 4+ points in their insomnia severity index at week 6, compared to 57.8% in the placebo group (p=0.02). Anxiety scores were significantly reduced compared to placebo (p<0.05), with a more pronounced effect among women (p<0.01). Additionally, subjective night sweat severity decreased, and objective measures of sleep duration were increased in the 5 Billion CFU cohort compared to placebo (p<0.05). Finally, the 5 Billion CFU cohort exhibited an increase in urinary GABA compared to placebo during the first week of use (p<0.05) and urinary GABA was inversely correlated with insomnia and anxiety scores at week 2 (r^2^=-0.30 and −0.48, respectively). All adverse events potentially related to product use were mild. A daily probiotic containing 5 billion CFU Lp815 significantly improved subjective symptoms of insomnia and anxiety, as well as objective measures of sleep after 6 weeks. Elevation of systemic GABA may contribute to these effects.

**Short Abstract:** Gamma-aminobutyric acid, or GABA, the major inhibitory neurotransmitter, acts via the central nervous system and microbial production in the gut. Strains that augment gut GABA are novel targets for improving sleep and mood. We previously demonstrated that *Lactiplantibacillus plantarum* Lp815 improves anxiety symptoms. The current study investigates Lp815’s impact on subjective sleep, mood, digestion and objective wearable sleep data over 6 weeks in adults with self-reported sleep disturbance. The trial was decentralized, double-blind, randomized and placebo-controlled. 139 Individuals (5 Billion CFU/day n=70, placebo n=69) were evaluated, aged 44 ± 13 years, 51.1% female and 55.4% Caucasian. A sub cohort (n=17, 5 Billion CFU n=9, Placebo n=8) collected urine samples for neurotransmitter analysis. Lp815 significantly increased sleep duration metrics, reduced night sweats, lowered insomnia severity index and lowered anxiety scores at 6 weeks compared to placebo (p<0.05), with greater anxiety reduction among women (p<0.01). Finally, urinary GABA increased during week 1 in the Lp815 cohort (p<0.05) and was inversely correlated with insomnia and anxiety. A daily probiotic containing 5 billion CFU Lp815 elevates systemic GABA and improves anxiety, subjective and objective measures of sleep after 6 weeks.

## Introduction

Gamma-aminobutyric acid (GABA), the major inhibitory neurotransmitter in the human nervous system, plays a pivotal role in the regulation of sleep and mood. Deficits in GABA can lead to a cascade of disorders ranging from anxiety and stress to mood swings and depression^1^. Investigations into the role of the microbiome in the Gut-Brain Axis (GBA) have highlighted its untapped potential in regulating mood, stress, and sleep via neurotransmitter production^2,3^.

Several bacterial strains, including lactic acid bacteria and bifidobacteria, can produce GABA^4^. However, the magnitude of GABA output varies among strains and the dynamics of their GABA synthesis within the human gastrointestinal environment are uncertain. This stems from the fact that GABA synthesis, typically a pH-stress response, stalls in less acidic environments^5,6^. We identified *Lactiplantibacillus plantarum* Lp815 for its elevated GABA production capability, finding that it synthesizes a significant amount of GABA at pH 4-6^7^, which includes much of the physiological range of the large intestine (∼ pH 5-7)^8–10^. If Lp815 indeed produces GABA within the human digestive tract, it may augment endogenous GABA levels or provide a natural alternative to GABAergic interventions commonly used for sleep, mood regulation, and pain management^11^. Due to GABA’s short plasma half-life and rapid clearance^11^, sustained production by a probiotic like Lp815 may offer a more stable and physiologically relevant delivery method. Indeed, our recent clinical trial found that Lp815 significantly improved symptoms of anxiety in anxious adults^12^. In the prior study, Lp815 also trended toward dose-dependent reduction in insomnia severity index (ISI) score, despite the cohort’s selection for chronic anxiety rather than sleep disturbance^12^.

Potential impacts on sleep are particularly important given the prevalence of sleep loss in the U.S. population. Gallup Poll estimates that per night, as of 2023, 20% of adults sleep 5 hours or less, 53% sleep 6 to 7 hours, only 26% sleep eight or more hours, and that deficits are more pronounced and impactful for women^13,14^. Sleep loss and the associated circadian (daily rhythm) disruption contribute to the development and progression of every major disease, including cognitive decline^15,16^, disordered mood^17–19^, metabolic illness^20^, cancer^21,22^, fertility deficits^23^, in-utero and childhood developmental disorders^24–26^. Sleep deficits are attributed to numerous factors, including the expansion of digital electronic devices into everyday life, demanding round-the-clock attention^27^, and use of these devices by young people during periods of developmental sensitivity for the sleep and circadian systems^28,29^.

Unfortunately, prescription medications with substantial risk profiles are common front-line treatments for symptoms of sleep disruption and associated anxiety. For example, the sleep medication Ambien (Zolpidem) issued a warning for dosing in women, calling for dosage to be lowered due to a sex difference in drug metabolism associated with increased side effects ^30,31^. Chronic use of prescription sleep aids is associated with earlier onset of cognitive decline ^32,33^. Similarly, when people are treated for anxiety (potentially comorbid with sleep disturbance in more than 70% of cases^34,35^), treatments frequently include mood modulators like selective-serotonin reuptake inhibitors (SSRIs) that have side effects including sexual dysfunction, weight gain and, unfortunately, sleep disturbance^36–38^. Over-the-counter sleep aids also have risks associated with chronic use, including circadian and hormonal disruption with melatonin^39–41^ (which is only available via prescription in many countries)^42^, and morning grogginess with melatonin^43^ and antihistamines^44,45^. Although behavioral interventions for disrupted sleep are effective, such as cognitive behavioral therapy for insomnia (CBTi)^46^, recent studies suggest that dropout rates during treatment are typically 14-40% ^47,48^. Alternative treatments are clearly needed to address the crisis in modern sleep.

This present study sought to determine if 5 Billion CFU daily of Lp815 would improve subjective and objective measures of sleep in a population with sleep disturbance. Additionally, the study evaluated the impact of Lp815 on anxiety, gastrointestinal symptoms, overall mood, severity of night sweats (if present) and quality of life. In a subset of participants, we tested the hypothesis that consumption of Lp815 elevates systemic GABA levels and that such elevations predict improvement in symptoms. Such preliminary evidence would support whether or not Lp815’s mechanism of action may be *in vivo* GABA production. The investigation was structured as a randomized, placebo-controlled, double-blinded, decentralized trial.

## Methods

### Eligibility

Participants were eligible if they met the following inclusion criteria: i) aged at least 18 years old, ii) ISI score of ≥ 15, corresponding to at least moderate symptoms, with at least moderate difficulty falling asleep, iii) if taking prescription medications for sleep and/or anxiety (e.g. Benzodiazepines, zolpidem, zaleplon, SSRIs, Buspirone, Tricyclics, MAOIs), or another class of medication for sleep, participants were required to maintain a stable dose for at least 4 weeks prior to randomization and throughout the study, iv) do a 4-week washout from any systemic anti-, pre-, pro-, or post-biotics prior to randomization, v) do a 4-week washout of any over-the-counter sleep aids (e.g., melatonin, valerian root) prior to randomization and refrain from use for the duration of the study.

Participants were also required to meet several lifestyle and behavioral inclusion criteria, including general good health at screening (Investigator discretion), English fluency, ability to provide informed consent, use a personal smartphone, receive the study product and wearable device at an address in the USA, complete study assessments over the course of up to 7 weeks, and wear an Oura ring sleep monitoring device daily.

Participants were excluded if they were taking prescription medication for anxiety and/or sleep (e.g., Benzodiazepines, Zolpidem, Zaleplon, SSRIs, Buspirone, Tricyclics, MAOIs) not on a stable dose, or for any irregular use of cannabis products. Participants were also excluded if they were taking any other prescription medications that impact systemic GABA levels (e.g., GABA supplements, barbiturates, gabapentin, pregabalin, valproate, tiagabine, vigabatrin, baclofen), or if they had received any investigational treatments within 30 days prior to randomization. Participants receiving Cognitive Behavioral Therapy for Insomnia (CBTi) were excluded. The following diagnoses and co-morbidities were excluded: a) Sleep Apnea that was not treated or well controlled, b) Inflammatory Bowel Disease (Crohn’s Disease or Ulcerative Colitis) or Irritable Bowel Syndrome, c) Narcolepsy, Restless Leg Syndrome, Circadian Rhythm Disorders, d) Major depressive disorder or bipolar disorder e) current or prior history of psychotic disorder, f) current diagnosis of Alcohol or Substance Abuse Disorder, g) presence of any infection or illness that causes chronic night-waking or impairs ability to retain urine overnight (e.g., chronic pain, uncontrolled benign prostatic hyperplasia, prostatitis, urinary tract infection, catheterization).

Participants with a known hypersensitivity or previous allergic reaction to rice maltodextrin were excluded as well as women who were currently pregnant, planning to become pregnant in the next 2 months, or breastfeeding. Participants with known external causes of sleep loss, such as a new baby, shift work, or international travel during the planned study period were excluded. Final exclusion was at the investigators’ discretion.

### Study Design

Participants completed up to a 16-week study consisting of screening period, randomization and shipping period, a baseline period and a 6-week product/placebo use period (**See Table 1**). Participants were randomized to one of two groups (1) GABA Probiotic Lp815 (5 Billion CFU/dose/day), and (2) matching placebo capsule. The investigators, study team and participants were blind to group assignment. Participants received the probiotic study product or placebo and study supplies after randomization. Demographic and medical history data, including concomitant medications, were collected during the screening period. Survey data, as described below, were collected during baseline and the study product/placebo use period. Additionally, participants were sent a validated wearable ring device^49–54^, the Gen3 Oura Ring (Oura Health OY, Oulu, Finland) to collect objective sleep data from baseline through the end of the study product/placebo use period. 20 Participants (10 per study group) were consented to participate in a sub study that required collection of urine at home at 7 timepoints during the study period (Baseline, Day 2, 4, 7, 14, 28, and 42 of the study product/placebo use period). All subjective data were collected through a mobile research app, the Consumer Health Learning & Organizing Ecosystem (Chloe) (People Science, Inc., Los Angeles, CA, USA), described in the Data Management System section.

**Table 1.** Demographics.

| Category | Total % (Count) |
| --- | --- |
| **Sex** |  |
| Female | 51.1 (71) |
| Male | 48.9 (68) |
| **Mean Age (STD)** | 44.1 (12.8) |
| **Ethnicity** |  |
| Caucasian | 55.4 (77) |
| African American | 15.1 (21) |
| Asian | 14.4 (20) |
| Hispanic | 8.63 (12) |
| Other | 6.48 (9) |

### Recruitment, Consent, and Enrollment

Participants were recruited through social media channels and researcher networks. Recruitment outreach consisted of IRB-approved advertising by email, digital marketing channels and word of mouth. The IRB-approved study landing page hosted on the People Science website led to an IRB-approved pre-screening questionnaire to determine individual qualification. Qualified participants were invited to download the Chloe mobile application and create an account to access the informed consent form, provide medical and social history and concomitant medications. Virtual electronic informed consent, including a study specific privacy authorization and the California Experimental Subject’s Bill of Rights (as applicable) were provided through the HIPAA-compliant cloud-based platform Chloe. Eligibility was determined by the investigators and study team after review of participant history and medications. Eligible participants who provided virtual electronic consent were automatically registered into the study by the platform.

### Data Management System

All data was securely stored on Chloe Amazon Web Services HIPAA-compliant servers. The Chloe platform contains modules for building and managing surveys, study landing pages, marketing outreach with tracking tools for recruitment, audited electronic consent forms, data management and analytics using an integrated relational database. Additionally, the platform contains a user-facing application for participants that delivers surveys, study instructions, calendar reminders, communication with study team, and personal data reports at study culmination. Data from completed assessments was automatically collected for analysis.

### Study Timeline and Activities

Participants’ activities were as follows (**See Supplemental Tables 1 and 2**). Participants gave informed consent and continued on to the Screening period where they completed demographics, medical history, concomitant medications and the Insomnia Severity Index (ISI)^55^. Eligible participants were randomized to GABA Probiotic Lp815 (5 billion CFU/capsule/day) or matching placebo and were shipped the study product or placebo and the Oura wearable ring device. All randomized participants confirmed shipment receipt before proceeding to the baseline period. Participants completed a 1-week baseline as described in **Supplemental Table 1**, including the ISI, the Generalized Anxiety Disorder 7-Item Questionnaire (GAD-7), the Gastrointestinal Symptom Rating Scale (GSRS), use of the Oura wearable ring device, daily survey questions regarding sleep, night sweats, mood and quality of life. Baseline was followed by six weeks of product or placebo use. The ISI, GAD-7 and GSRS were repeated at bi-weekly intervals. Daily surveys with an additional question on product use adherence, deviation from normal routine and Oura ring use continued throughout the study. Participants were prompted to report any adverse events (AEs) weekly or advised to reach out to the study team at any other time to report an AE. A study experience survey and participant impression of change question were administered at the end of study.

### Sub Study Urine Collection and Analysis

Urinary neurotransmitter levels (GABA, serotonin, dopamine, norepinephrine, epinephrine, glutamate, glycine, histamine and Phenethylamine (PEA) were assessed in sub study participants at baseline, day 2, 4, 7, 14, 28 and 42 of product use (**See Supplemental Table 2**). Briefly, participants collected first morning urine samples at home and shipped samples for analysis (Doctor’s Data, St. Charles, IL, USA). As reference ranges varied across samples, data were normalized to the reference ranges provided for each sample and are presented as percentiles within range. Participants whose baseline data fell below range or above range at baseline were excluded from analysis (n=2).

### Data Evaluability, Analysis and Statistics

Participants were required to fill out their baseline ISI survey, at least 2 out of 3 ISI surveys during the study product/placebo use period and to not miss a consecutive week or more of study product/placebo use during the study period. Participants were withdrawn from the study if they needed to take antibiotics or new sleep medications during the product use window, as antibiotics disrupt the gut microbiome and as sleep medications would confound study results. Individual days of data were excluded from daily metric analysis if product/placebo consumption was missed. These exclusions were made prior to unblinding. All analyses were conducted using Python (version 3.9.13) Jupyter Notebooks (Project Jupyter, Beaverton, OR, USA). Normality was assessed using the Shapiro-Wilk (SW) test. Non-parametric statistics (Mann-Whitney U, MW) were used in cases where data were not normally distributed. Holm-Bonferroni correction was used in the case of multiple comparison testing, alongside general linear mixed models (GLMM) for evaluating impacts of group, demographics and time. The Gamma family was used as data were positive, continuous and right skewed, with variance increasing with the mean. Mann-Kendall (MK) tests were used to evaluate potential trends over time in daily data. Student’s T-tests were used to evaluate differences between normally distributed measures with a Mixed Linear Model (MLM) in the case of evaluating demographic impacts on normally-distributed anxiety data. Fisher’s Exact Tests (FET) were used to evaluate differences in percent improvers.

## Results

### Recruitment and Conduct

This study was approved by Sterling Institutional Review Board (12481-NACraft) and registered with ClinicalTrials.gov (NCT06789718). All participants gave informed consent. All study procedures were conducted by People Science following Good Clinical Practice (GCP) guidelines and in accordance with the ethical principles of the Declaration of Helsinki and its amendments. Informed consent was obtained from all participants.

### Demographics and Sample Size

Of 150 participants who were randomized, N=139 were evaluated (Placebo=69, 5 Billion CFU=70). Eleven were withdrawn due to: antibiotic use (3), participant decision (1), lost to follow-up (6) and use of exclusionary medication not reported at screening (1). One participant used antibiotics in the last week of the study but their data prior to antibiotic use was included, as they met data evaluability criteria described above in the Data Evaluability methods. The cohort was 51.1% female (n=71), 48.9% male (n=68). Mean age was 44.1 ± standard deviation of 12.8 years. The cohort was 55.4% Caucasian (n=77), 15.1% African American (n=21), 14.4% Asian American (n=20), 8.63% Hispanic (n=12), and 6.48% Other (n=9) (**Table 1**). Participant medications are described in **Supplemental Table 3**.

### Insomnia Severity Index

ISI score did not differ significantly between groups at baseline and decreased across the study period in both groups (MW p<0.05). ISI score decreased significantly compared to placebo at week 6 (ISI 9.31 ± 4.58 in 5 Billion CFU vs. 12.0 ± 5.78 in Placebo, MW p=0.01). The percentage of individuals who improved by at least 4 points was greater in 5 Billion CFU (77.3%) than Placebo (57.8%) (FET p=0.02). Improvement was spread across ISI questions, with statistical improvements in difficulty falling asleep (MW p=0.01), difficulty staying asleep (MW p=0.04) and problems waking up too early (MW p=0.03). Increased satisfaction with sleep pattern and decreased worry about sleep problems trended strongly toward improvement (MW p=0.05 for both). (See **Table 2**. Of note, “Extent of Interference With Daily Function” (MW p=0.42) was not statistically significant between in either group). Cross-sectional analysis of individuals who began with at least mild insomnia (5 Billion CFU=64, Placebo=63), moderate insomnia (5 Billion CFU = 48, Placebo=48) or severe insomnia (5 Billion CFU=10, Placebo=11) revealed that reductions in 5 Billion CFU compared to Placebo were statistically significant in all cases (MW p=0.01 for all), with increased severity corresponding to a greater drop in score (See **Tables 3 and 4**). Additionally, those with severe baseline insomnia exhibited a statistically significant drop by week 4 (MW p=0.04, −11.1 ± 5.7 vs −6.9 ± 4.8).

**Table 2.** Improvement in Insomnia Severity Index by Question. Insomnia Severity Index mean (standard deviation) score at week six by question. Bold text highlights questions with a statistically significant group difference. ^#^Note that a lower number indicates greater satisfaction with the current sleep pattern in question 4. Lower scores indicate better outcomes for all questions.

| Question | p-value | 5B CFU Score | Placebo Score |
| --- | --- | --- | --- |
| **1. Difficulty Falling Asleep** | **0.01** | **1.1 (0.80)** | **1.5 (0.82)** |
| **2. Difficulty Staying Asleep** | **0.04** | **1.4 (0.82)** | **1.8 (0.98)** |
| **3. Problem Waking Up Too Early** | **0.03** | **1.2 (0.92)** | **1.6 (1.2)** |
| 4. Satisfaction With Current Sleep Pattern<sup>#</sup> | 0.05 | 1.9 (1.0) | 2.3 (0.98) |
| 5. Extent of Interference With Daily Function | 0.42 | 1.1 (1.0) | 1.3 (1.2) |
| 6. How Noticeable is the Sleep Problem in Terms of Impairing Quality of Life? | 0.11 | 1.4 (1.1) | 1.6 (1.1) |
| 7. Worry or Distress About the Current Sleep Problem | 0.05 | 1.3 (0.92) | 1.7 (1.1) |

**Table 3.** Magnitude of Drop in ISI at Week 6 Increases with Baseline Insomnia Severity in 5 Billion CFU Cohort. Magnitude of ISI reduction at week 6 stratified by starting insomnia severity. *Indicates significant difference between the 5 Billion CFU time point compared to placebo.

| <u>Week 6</u> | <u>Mild ISI +</u> | <u>Moderate ISI +</u> | <u>Severe ISI</u> |
| --- | --- | --- | --- |
| <u>5 Billion CFU</u> | -6.97 ± 5.48* | -8.67 ± 5.09* | -12.50 ± 5.94* |
| <u>Placebo</u> | -5.26 ± 4.71 | -5.67 ± 4.86 | -6.5 ± 5.36 |

**Table 4.** ISI Total Scores. Total ISI score at baseline and by week of intervention mean ± SD.* Indicates significant difference between the 5 Billion CFU time point compared to placebo.

|  | <u>Baseline</u> | <u>Week 2</u> | <u>Week 4</u> | <u>Week 6</u> |
| --- | --- | --- | --- | --- |
| **<u>5 Billion CFU</u>** | 16.23±4.41 | 12.06 ± 4.71 | 10.21 ± 4.69 | **9.31 ± 4.58\*** |
| **<u>Placebo</u>** | 17.21 ± 4.53 | 12.84 ± 5.07 | 11.81 ± 5.51 | 12.00 ± 5.78 |

### Anxiety Symptoms

Within-individual reduction in GAD-7 score was of significantly greater magnitude in the 5 Billion CFU cohort at weeks 2, 4 and 6 compared to placebo (Student’s p<0.01) (See **Table 5**). Additionally, women in the 5 Billion CFU cohort exhibited statistically greater decreases in GAD-7 score compared to men (MLM r^2^=0.1, F=3.6, p=0.001, men averaging +1.7 points). All groups decreased GAD-7 score and raw scores did not differ (See **Supplemental Figure 1**). Improvement did not differ statistically by question (data not shown). Finally, groups did not differ in the proportion of participants improving their GAD-7 total score by 4+ points at 6 weeks, with 37.5% of placebo and 50.9% of 5 billion CFU improving more than one category (e.g., going from moderate to minimal/no anxiety).

**Table 5.** Change in Total GAD-7 Anxiety Score from Baseline by Week of Product Use. GAD-7 Anxiety score mean change ± SD at weeks 2, 4 and 6 of intervention.* Indicates significant difference between the 5 Billion CFU time point compared to placebo.

|  | **<u>Week 2</u>** | **<u>Week 4</u>** | **<u>Week 6</u>** |
| --- | --- | --- | --- |
| **<u>5 Billion CFU</u>** | -3.54 ± 3.92* | -4.20 ± 4.02* | -4.85 ± 3.62* |
| **<u>Placebo</u>** | -2.96 ± 4.40 | -4.00 ± 4.50 | -3.98 ± 4.14 |

### Daily Mood, Subjective Sleep Symptoms, and Quality of Life

Night sweats trended down across the study period in 5 Billion CFU (MK p<0.05), with women exhibiting a steeper downward slope compared to men (MK p<0.05) (See **Figure 2** and **Supplemental Figure 2**). Sleep quality and daily mood trended up comparably in both groups (MK p<0.05, no group difference in trend over time). No trend over time or group differences were observed for subjective frequency of mid-sleep wakeups or self-reported bedtime. However, participants in the 5 Billion CFU group exhibited greater night sweat intensity (2.48 out of 5 vs 1.5 out of 5) at study onset compared to placebo (Student’s p<0.05).

**Figure 1.**
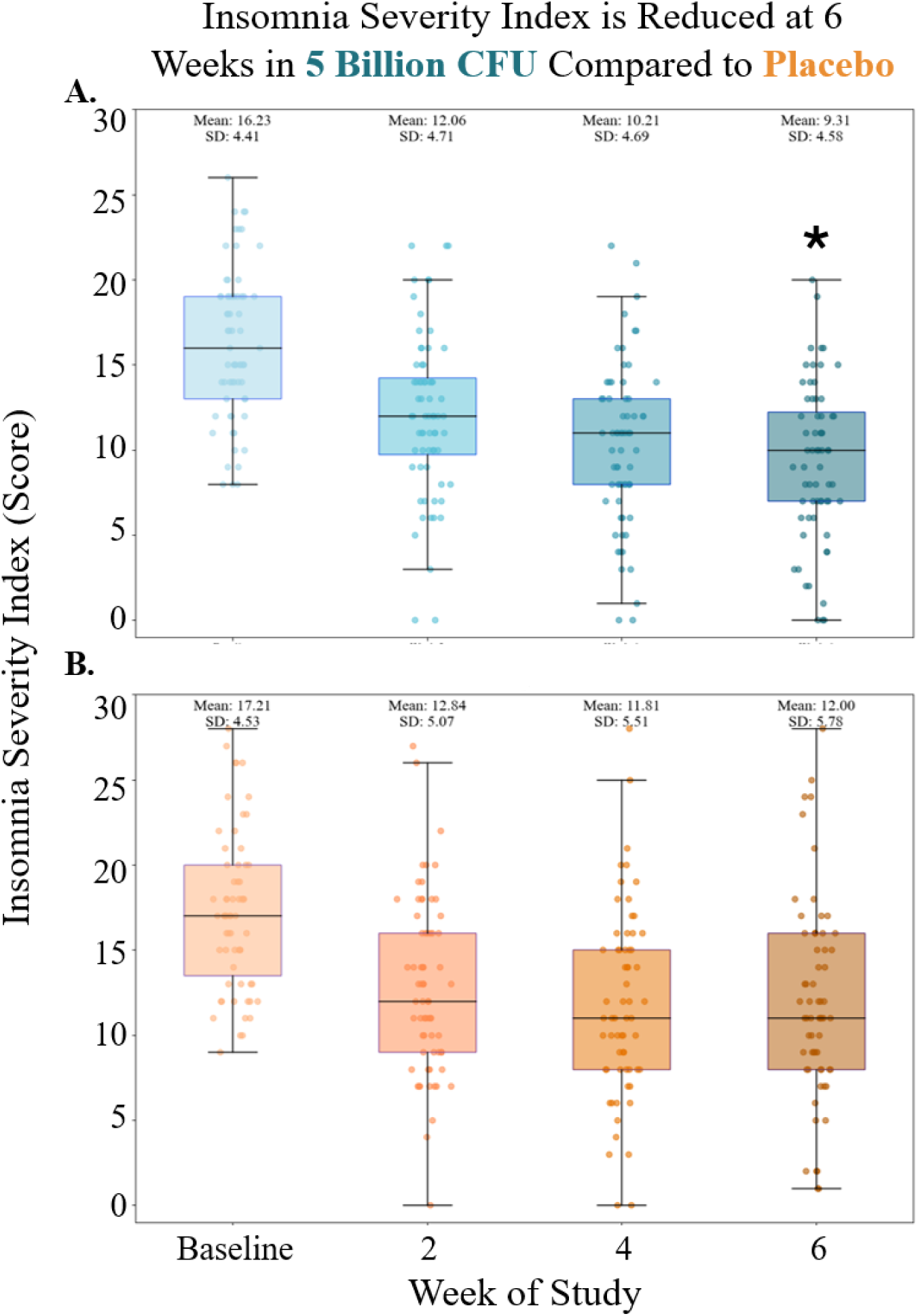
Insomnia Severity Index is Reduced at 6 Weeks in 5 Billion CFU Compared to Placebo. Box plots are overlaid with dots representing individuals’ ISI scores at baseline, Week 2, Week 4 and Week 6. Means and SDs above upper whiskers represent change for that group at that time point. 5 Billion CFU is depicted in blue (A) and placebo in orange (B).

**Figure 2.**
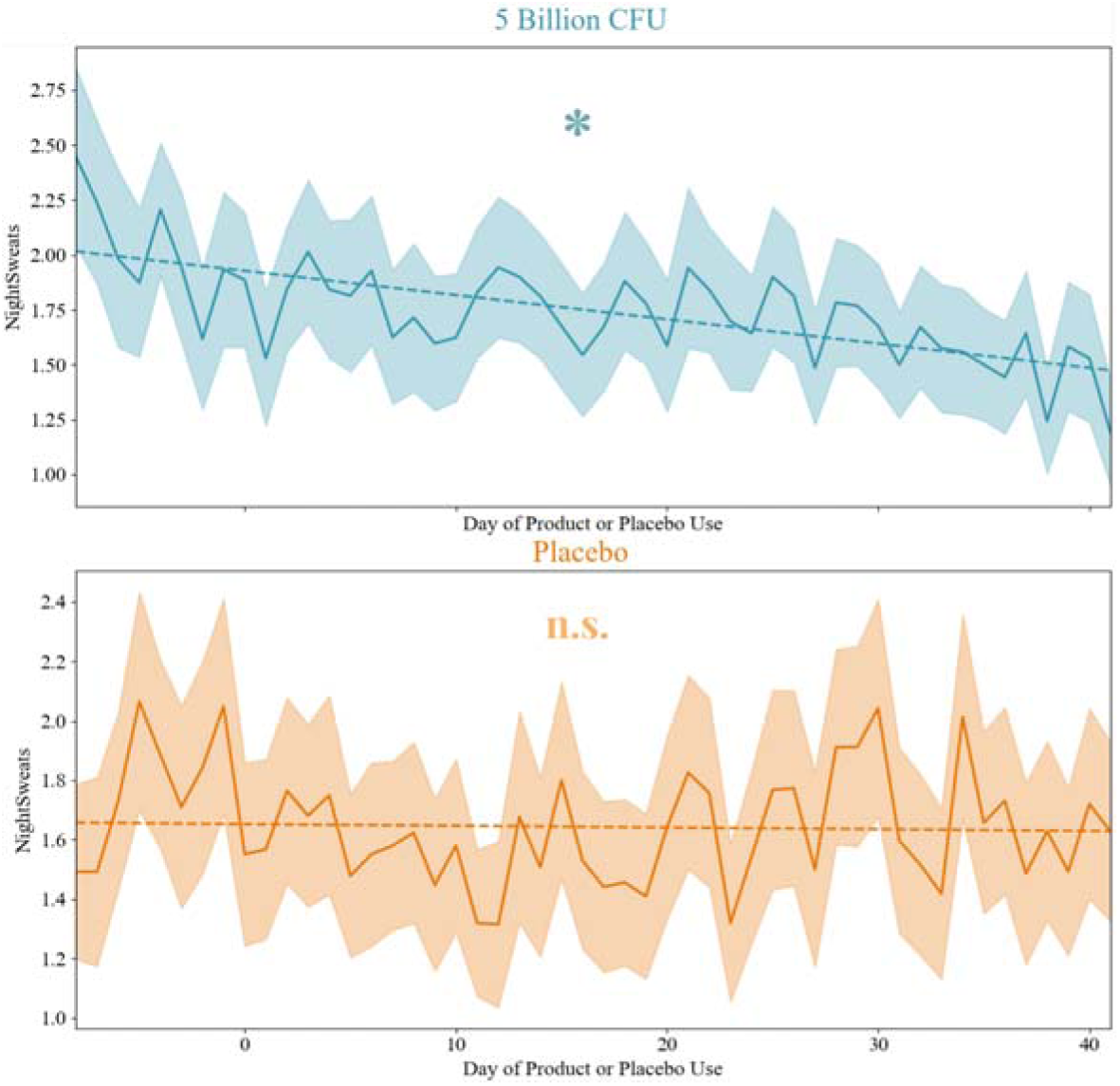
Night Sweats Decrease Significantly in 5 Billion CFU. Mean ± SEM of daily rating of night sweat severity (0-5) in 5 Billion CFU (top, blue) and placebo (bottom, orange).

### Gastrointestinal Symptom Rating Scale (GSRS)

GSRS did not differ at baseline or end of study, between or within groups (**See Table 6**). Total GSRS score ranged from 1.66 ± 0.11 to 2.23 ± 0.12 across the study, corresponding to very mild gastrointestinal symptoms.

**Table 6.** Gastrointestinal Symptom Rating Scale GSRS Subscale and total weighted average score means (SD) at baseline and end of study.

| Group | Time point | Reflux Score | Abdominal Pain Score | Indigestion Score | Diarrhea Score | Constipation Score | Total GSRS Score |
| --- | --- | --- | --- | --- | --- | --- | --- |
| 5B CFU | Baseline | 2.12 (0.16) | 2.21 (0.14) | 2.53 (0.17) | 1.91 (0.16) | 2.25 (0.18) | 2.20 (0.13) |
| Placebo | Baseline | 2.12 (0.17) | 2.15 (0.14) | 2.49 (0.15) | 2.06 (0.15) | 2.34 (0.16) | 2.23 (0.12) |
| 5B CFU | End of Study | 1.68 (0.13) | 1.66 (0.10) | 1.87 (0.10) | 1.52 (0.11) | 1.78 (0.13) | 1.70 (0.08) |
| Placebo | End of Study | 1.66 (0.11) | 1.76 (0.12) | 2.12 (0.15) | 1.72 (0.13) | 1.82 (0.13) | 1.82 (0.10) |

### Wearable Sleep Metrics

Sleep metrics did not differ at baseline and first week of product us (GLMM p>0.05). Time in bed (**Figure 3A)**, total sleep duration (**Figure 3B**), deep sleep duration (**Figure 3C**) and light sleep duration (**Figure 3D**) improved in the 5 Billion CFU cohort. Time in bed was significantly greater for 5 Billion CFU from Week 1 to end of study during product us (GLMM group effect p=0.005, Holm-Bonferroni MW corrected difference p=0.0004, 9 minutes). Total sleep duration was greater in the 5 Billion CFU cohort (GLMM group difference p=0.005, Holm-Bonferroni MW p=0.03, 6 minutes). Deep sleep and light sleep duration were also greater with 5 Billion CFU (Deep Sleep GLMM group difference p=0.002, Holm-Bonferroni MW p<1*10^-4^), Light Sleep GLMM group difference p=0.005, Holm-Bonferroni MW p=0.18). Cardiovascular metrics differed by group at baseline, with placebo exhibiting lower nighttime average heart rate, lower breathing rate and higher heart rate variability (See **Supplemental Figure 3**). Because these factors were present at baseline and did not differ by phase of study, they were not considered related to the intervention.

**Figure 3.**
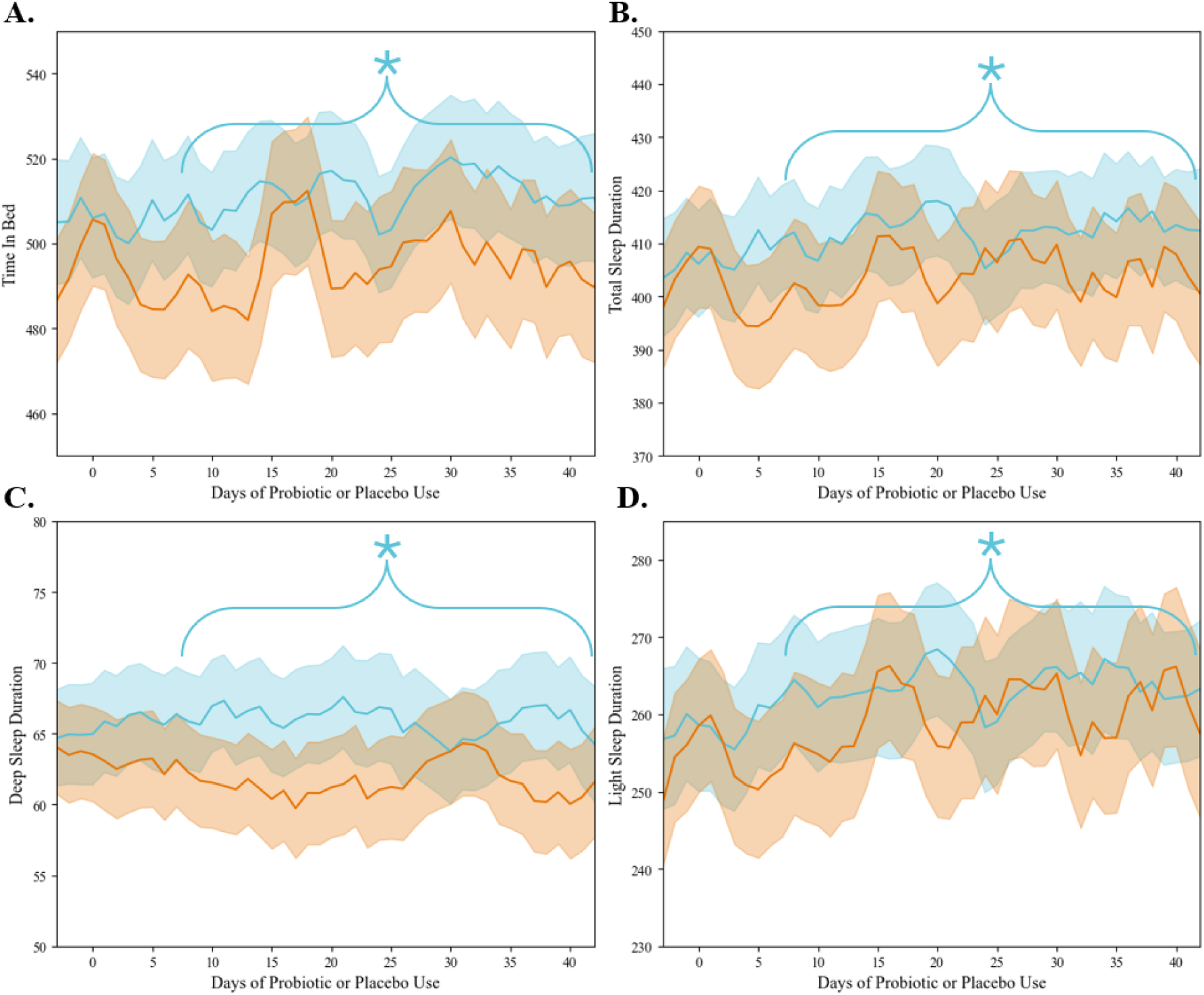
5 Billion CFU Exhibits Longer Time in Bed, Total Sleep Duration, Deep and Light Sleep Duration. Mean ± SEM of time in bed (A), total sleep duration (B), deep sleep duration (C) and light sleep duration (D) in 5 Billion CFU (blue) and Placebo (orange). Star indicates significant difference between groups over the bracketed time window, day 7-42 of intervention. Measures did not differ at baseline and first week of product use.

### Sub Cohort Analysis of Urinary Neurotransmitters

Urinary GABA values differed after 7 days of supplementation, with 5 Billion CFU at 67th percentile of normal range (±32) and placebo at 32nd percentile (±14), (Welch’s t-test p=0.04). Other days trended toward higher levels in 5 Billion CFU but did not differ statistically (See **Figure 4A**). Placebo had only two participants per time point after day 14, so only 5 Billion CFU data is shown in (**Figure 4B**). For data from all participants see (**Supplemental Figure 4**). Urinary GABA increased in 5 Billion CFU across the study period (Mann-Kendall p=0.01), from 45th (±16%) to 70th percentile (±17%) of normal range (See **Figure 4B**), remaining relatively stable from end of Week 1 to Week 6. Additionally, higher ISI and GAD-7 scores were associated with lower GABA levels at Week 2 (r^2^=0.30 and 0.38. respectively) but not at baseline (See **Figure 4C-F**.). Similar trajectories of improvement were observed across time in ISI, GAD-7 and night sweats as in the overall cohort (See **Supplemental Figures 5-7**). Histamine was higher in the placebo group at baseline and throughout the study (MW p<0.05), although neither group exhibited a trend in concentration across the study (**See Supplemental Figure 8).** Other neurotransmitters did not differ statistically by group (See **Supplemental Figure 8**), although glutamate exhibited an increasing trend in the 5 Billion CFU sub cohort (See **Supplemental Figure 8B**).

**Figure 4.**
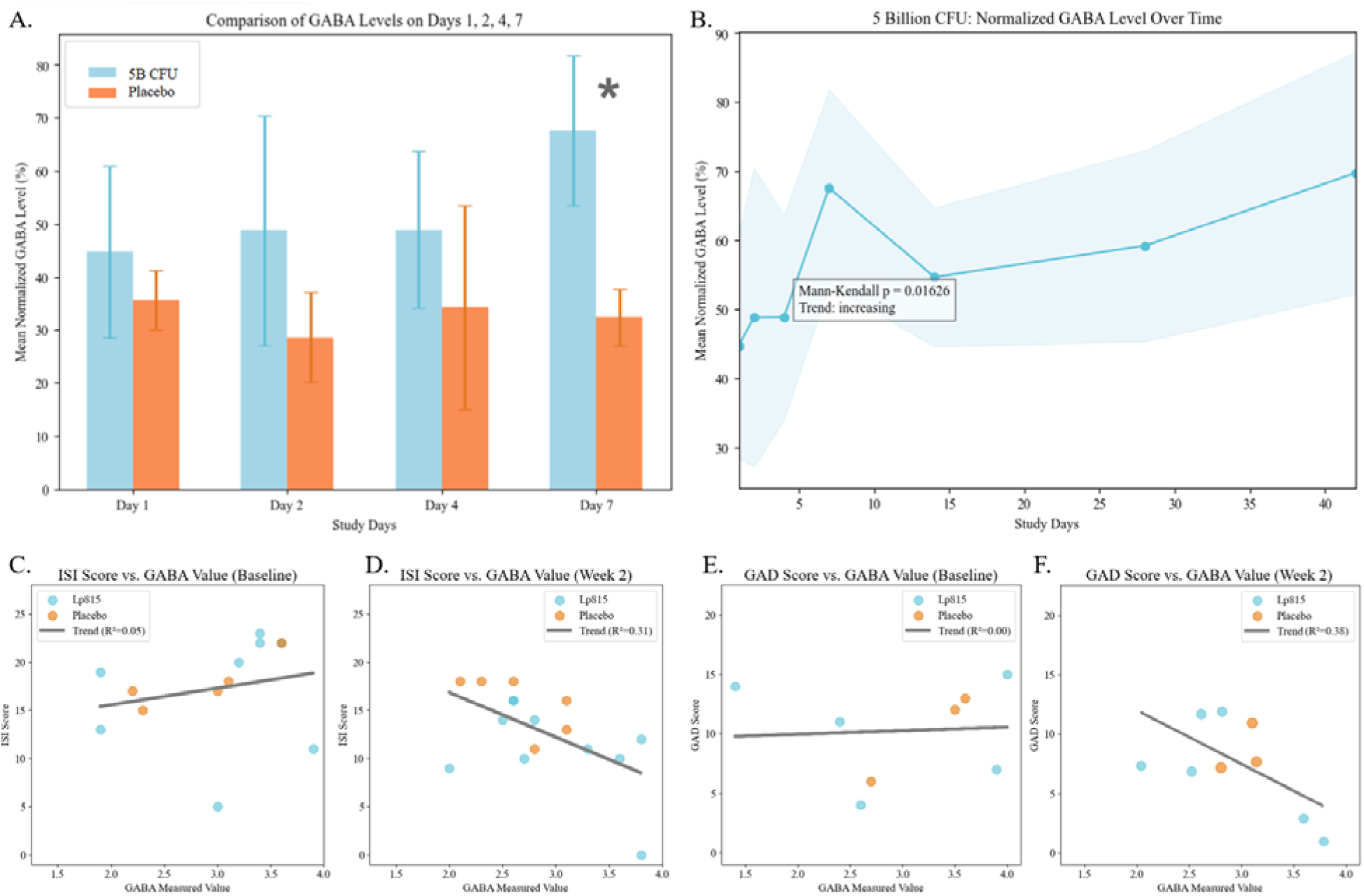
Urinary GABA Increases with 5 Billion CFU Lp815 After 1 Week and is Associated with Reduced Insomnia and Anxiety at Week 2. A) Mean ± SEM of reference range-normalized GABA levels in 5 Billion CFU (blue) and placebo (orange) across the first week of supplementation. B) 5 Billion CFU Mean ± shaded SEM GABA levels increase rapidly in week 1 and are relatively stable to week 6 (data from Placebo not available past week 1). C-D) Scatter of GABA baseline values and ISI score at baseline (C) shows no correlation, whereas week two shows decreasing ISI score with increasing GABA (D). E) Scatter of GABA baseline values and GAD-7 score at baseline (E) shows no correlation. Comparably to ISI score, GABA and week 2 GAD-7 values are inversely correlated (F). Blue dots represent data from Lp815 supplemented participants, and orange dots represent natural variation within placebo participants.

### Adverse Events

Six mild AEs were deemed possibly related to study participation. Three occurred in 5 Billion CFU, with an instance each of heartburn, diarrhea and stomach pain. Three AEs: diarrhea, bloating and stomach pain were reported in the placebo group. All these AEs were mild and resolved. Additionally, n=22 reported AEs were deemed unlikely to be related to study participation (e.g., post-vaccination malaise, colds and flu, headache and joint pain; data not shown). One serious AE occurred but was not considered attributable to the intervention: a relapse in a participant with a history of serotonin syndrome in the 5 Billion CFU cohort. This participant was briefly hospitalized, recovered and was withdrawn.

## Discussion and Conclusions

The present study demonstrates that consumption of Lp815, a probiotic strain known to produce GABA under physiological conditions^7^, is associated with a significant and meaningful reduction in subjective and objective measurements of sleep disturbance and anxiety after 6 weeks of supplementation compared to placebo. These improvements included insomnia severity score (ISI), anxiety score (GAD-7), night sweat severity, time in bed, total sleep duration and sleep architecture (deep sleep and light sleep duration). Interestingly, reduction in anxiety was greater among women and participants with more severe baseline insomnia symptoms experienced earlier and more pronounced improvements in ISI score. Further, our preliminary sub study suggests that increased systemic GABA concentrations may be responsible for these benefits. GAD-7 and ISI scores were inversely correlated to GABA concentration within all participants after 2 weeks of product or placebo use. We also observed the same trajectory of improvement in ISI, GAD-7 and night sweats in these sub study participants. Of note, urinary GABA was elevated after 1 week of Lp815 consumption compared to placebo, consistent with the hypothesis that the probiotic increases gut-derived GABA, some of which may enter general circulation and persist at more stable concentrations than bolus-GABA from pharmaceuticals^11^. Interestingly, we observed relatively stable GABA levels in supplemented participants after the first week, and no evidence of tolerance (i.e., no reduction in intervention effects past that first week; on the contrary, metrics continued to improve throughout the study period). Moreover, although concentrations did not statistically differ between Lp815 and placebo, we observed a trend toward increasing glutamate in Lp815 over the first week of consumption. Such an elevation in both neurotransmitter concentrations would be consistent with the known interconversion of glutamate and GABA^56^. The probiotic was safe and well-tolerated by participants. Adverse events related to Lp815 use consisted of mild digestive disturbance which resolved during the study and occurred at the same rate in placebo. Similar to the placebo, the intervention did not interfere with daily function, suggesting Lp815 supplementation does not lead to the grogginess or cognitive impairment that has been associated with prescription^36–38^ and over-the-counter sleep aids^39–41^.

These results extend previous findings on the positive impact of neurotransmitter or tryptophan modulating strains on sleep and mood. Rodent studies of *Lactiplantibacillus* LPB145 demonstrated antidepressive effects^57^, and a similar study in mice of *Bifidobacterium bifidum* TMC3115 found both reduced anxiety and increased intestinal GABA^58^. A number of small human clinical trials have shown the psychoactive effects of specific strains. Depressive symptoms dropped after 8 weeks of daily 30 Billion CFU *Lactiplantibacillus plantarum* PS128 supplementation in a small, open-label trial^59^. Although the study did not test biochemical markers, a previous study on PS128 found that its use can increase dopamine and serotonin levels in the brains of mice^60,61^. The same strain and dosage was later evaluated in 40 participants with self-reported insomnia in a double-blind, placebo-controlled trial, with outcomes including decreased depression, fatigue and fewer awakenings from deep sleep measured using polysomnography^62^. The largest previously published human study involved probiotic *Lactiplantibacillus plantarum* P8 and investigated 103 stressed adults. It was found that 12 weeks of 20 Billion CFU daily reduced subjective stress and anxiety compared to placebo, reduced pro-inflammatory cytokines IFN-_γ_ and TNF-_α_, improved memory and cognition^63^. Finally, our previous study of 83 adults^12^ demonstrated that a daily dose of 5 Billion CFU Lp815 significantly improved symptoms of anxiety compared to placebo in a population of anxious adults. Together, studies of model organisms to blinded, placebo-controlled trials demonstrate the ability of related probiotic strains to improve mood, sleep and stress. To our knowledge, the present study is the largest to date on this topic in humans, the first to assemble comprehensive subjective and objective measurements of sleep and mood in a real-world setting and the first to tie these findings to the hypothesized mechanism of action.

This study was a decentralized clinical trial, intentionally designed to collect data at home during the everyday lives of participants. Participants’ measurements were obtained using an app-based interface, use of wearable devices and at-home urine collection rather than in-person evaluation at a clinic. This design was chosen to evaluate the impacts of the probiotic under real-world conditions with minimal compromise on data quality^64–66^. However, due to the nature of real-world conditions (e.g., the fact that this trial was conducted over the holiday season), this dataset may exhibit greater variability due to within-individual differences and variable outside circumstances in comparison to traditional site-based clinical trials^64–66^. Indeed, it is possible that variability in response to this intervention would be lower at other times of year, as participants likely responded variably to the holidays (with some experiencing increased stress and poorer sleep due to travel and family life^67,68^, and some experiencing decreased stress and better sleep due to these same factors).

Individual response to the intervention appeared influenced by both biological and clinical baseline characteristics. Variability in GABA-ergic tone, as seen in some sub study participants with out-of-range baseline GABA levels, may contribute to differential effects. Additionally, greater baseline insomnia severity was associated with earlier and more pronounced improvements. These findings suggest that neurochemical and symptomatic starting points may shape responsiveness to GABA-producing probiotic supplementation. Future studies linking urinary or serum samples’ neurotransmitter levels with gut microbiome profile and intestinal pH will further clarify the relationships among Lp815 ingestion, host microbiota and suitability for GABA production and initial neurotransmitter status. Although our findings suggest a benefit to the large majority of people with symptoms of sleep disturbance, future studies will clarify the biological profile that most benefits from Lp815 supplementation.

Together, daily consumption of a 5 billion CFU probiotic containing Lp815 led to a significant and meaningful reduction in subjective and objective measures of sleep disturbance after 6 weeks, with trends toward improvement observed across the entire product use period. This dose resulted in more individuals categorically reducing insomnia severity compared to placebo. Additionally, our previous findings of reduced anxiety with 5 billion CFU daily appear to extend beyond anxious cohorts to those with sleep disturbance, with women exhibiting stronger improvements than men. Finally, preliminary evidence supports the hypothesis that Lp815 acts through increasing systemic GABA. The probiotic appears safe and well-tolerated, with no interference in daily functioning. The GABA-producing probiotic Lp815 may offer a safe and natural option to address symptoms of anxiety, night sweats, sleep disturbance, increase sleep duration and satisfaction in people with symptoms of insomnia.

## Author Contributions

Study conceptualization: NC, MCBE, ADG, PLO, AK, CJD, DK, NPZ. Data curation: PLO, ADG. Formal data analysis and visualization: ADG. Funding acquisition: MCBE, NC, NPZ. Investigation: MCBE, AK, NC, ADG, JM, VL, PLO. Methodology: NC, MCBE, ADG, PLO, CJD, DK, NPZ. Project administration: MCBE, AK, PLO, NC, JM, VL,

ADG. ADG prepared the manuscript with help from MCBE. All authors edited and approved the final manuscript.

## Acknowledgements

The authors would like to thank the Doctor’s Data team for the assay of urinary neurotransmitter levels throughout the study.

## Trial Registration

The trial was approved by Sterling IRB (12481-NACraft) and registered with ClinicalTrials.gov (NCT06789718)

## Data Availability

Data are available upon reasonable request from the corresponding author.

