## Supplemental Figures for "GABA Probiotic Lactiplantibacillus plantarum Lp815 Improves Sleep, Anxiety and Increases Urinary GABA: a Randomized, Double-Blind, Placebo-Controlled Study"

**Supplemental Materials**

**Supplemental Table 1. Study Activities**

| **Protocol Activities** | **Screening^(a)^** | **Randomize and Ship ^(b)^** | **Study Duration** | | | | | | |
| --- | --- | --- | --- | --- | --- | --- | --- | --- | --- |
|  |  |  | **Baseline** | **Study Product/Placebo Use Period**  **(6 weeks)** | | | | | |
|  |  |  |  | **Week 1** | **Week 2** | **Week 3** | **Week 4** | **Week 5** | **Week 6** |
|  | **Up to Day -28** | **Up to**  **Day 0** | **Day 1-7** | **Day 8-14** | **Day 15-21** | **Day 22- 28** | **Day 29-35** | **Day 36-42** | **Day 43-49** |
| Informed Consent | X |  |  |  |  |  |  |  |  |
| Demographics | X |  |  |  |  |  |  |  |  |
| Medical History | X |  |  |  |  |  |  |  |  |
| ISI^(c,d^**^)^** | X^(d^**^)^** |  | X |  | X |  | X |  | X |
| Eligibility Confirmation | X | X |  |  |  |  |  |  |  |
| Randomization^(e^**^)^** |  | X |  |  |  |  |  |  |  |
| Participant Receipt of Product and Supplies Shipment |  | X |  |  |  |  |  |  |  |
| GAD-7^(f^**^)^** |  | X | X |  | X |  | X |  | X |
| GSRS^(g^**^)^** |  |  | X |  |  | X |  |  | X |
| Use of Oura Ring wearable device^(h^**^)^** |  |  | DAILY | | | | | | |
| Survey questions^(i^**^)^** |  |  | DAILY | | | | | | |
| Study product/  placebo use^(j^**^)^** |  |  |  | DAILY | | | | | |
| Adverse Event Survey^(k^**^)^** |  |  |  | WEEKLY | | | | | |
| P-GIC^(l^**^)^** |  |  |  |  |  |  |  |  | X |
| Experience Survey |  |  |  |  |  |  |  |  | X |

a. Screening occurred within 4 weeks prior to Randomization
b. Randomization and Study Product/Supplies Shipping occurred within 3 weeks prior to Baseline.
c. Insomnia Severity Index
d. Screening ISI score ≥ 15, with question on difficulty falling asleep ≥ 2 (moderate)
e. Study participants were randomized to one of 2 groups: (1) *Lactiplantibacillus plantarum* Lp815 (5 Billion CFU/day), (2) Matching placebo
f. Generalized Anxiety Disorder - 7 Scale
g. Gastrointestinal Symptom Rating Scale
h. Use of a health tracking wearable device (Oura Ring) during sleep
i. Participants completed a morning survey to review objective data from wearable device collection and answer questions about their prior night’s sleep quality, daytime alertness, mood, severity of night sweats (0-5, 0= none, 5= most severe), quality of life and any deviations from regular routines (e.g., cannabis and alcohol use, concomitant medications)
j. Take the study product/placebo capsule 1x daily with food
k. Participants answered an Adverse event question weekly during the Study Product/Placebo Use Period
l. Patient Global Impression of Change

**Supplemental Table 2. Sub-Study Activities**

| **Protocol Activities** | **Randomize and Ship** | **Study Duration** | | | | | | |
| --- | --- | --- | --- | --- | --- | --- | --- | --- |
|  |  | **Baseline** | **Study Product/Placebo Use Period**  **(6 weeks)** | | | | | |
|  |  |  | **Week 1** | **Week 1** | **Week 1** | **Week 2** | **Week 4** | **Week 6** |
|  | **Up to**  **Day 0** | **Day 0** | **Day 2** | **Day 4** | **Day 7** | **Day 14** | **Day 28** | **Day 42** |
| Sub Study  Informed Consent | X |  |  |  |  |  |  |  |
| Neurotransmitter Panel Urine Test |  | X | X | X | X | X | X | X |

**Supplemental Table 3. Participant Medications**

| **Medication** | **Number of Participants** | **Purpose** |
| --- | --- | --- |
| **Atorvastatin** | **4** | **Lower Cholesterol** |
| **Pravastatin** | **1** | **Lower Cholesterol** |
| **Lisinopril** | **6** | **Hypertension** |
| **Amlodipine** | **2** | **Hypertension** |
| **Furosemide** | **1** | **Hypertension, Edema** |
| **HCTZ** | **1** | **Hypertension, Edema** |
| **Atenolol, Fenofibrate** | **1** | **Hypertension, Dyslipidemia** |
| **Cymbalta, Propranolol** | **1** | **Mood Disorder/Pain. Hypertension** |
| **Metoprolol, Escitalopram** | **1** | **Hypertension, Mood Disorder, Pain** |
| **Topiramate, Fluoxetine** | **1** | **Migraine or Epilepsy, Mood Disorder/Pain** |
| **Levothyroxin** | **4** | **Hypothyroid** |
| **Finasteride, Minoxidil** | **1** | **Hair Regrowth** |
| **Testosterone** | **1** | **Hormone Replacement Therapy** |
| **Oral Contraceptive (unspecified)** | **1** | **Pregnancy Prevention** |
| **Tadalafil** | **1** | **Benign Prostatic Hyperplasia** |
| **Empagliflozin, Glipizide** | **1** | **Type 2 Diabetes** |
| **Semaglutide, Esomeprazole** | **1** | **Weight Loss, Proton Pump Inhibitor** |
| **Advair** | **1** | **Allergy** |
| **Prilosec** | **1** | **Proton Pump Inhibitor** |

**Supplemental Figure 1. Raw GAD-7 Anxiety Scores Across Study**

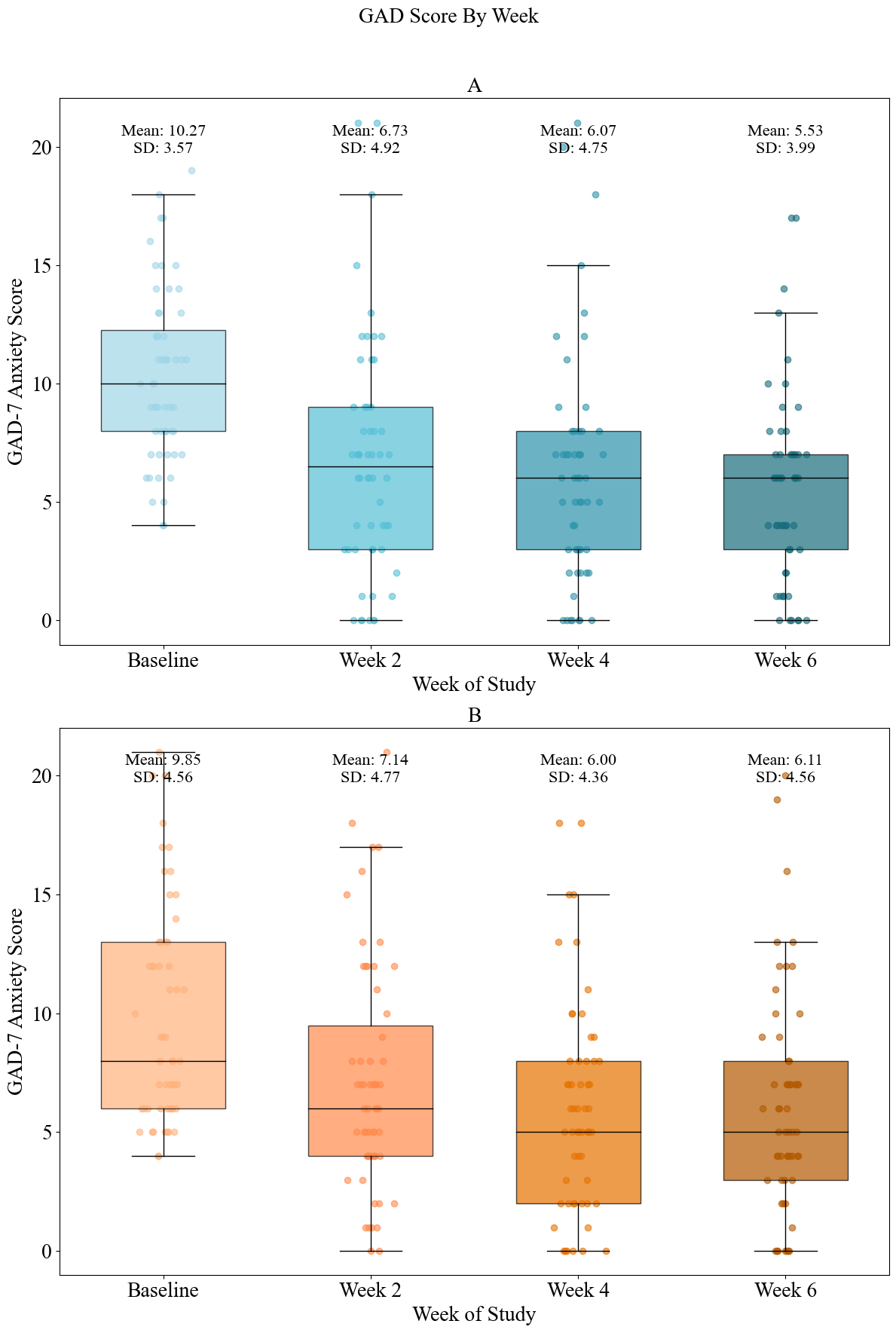

**Supplemental Figure 1. GAD-7 Raw Score Decreases Across Both Groups Over Six Weeks.** Box plots are overlaid with dots representing individuals’ GAD-7 scores at baseline, Week 2, Week 4 and Week 6. Means and SDs above upper whiskers represent change for that group at that time point. 5 Billion CFU is depicted in blue (top) and placebo in orange (bottom).

**Supplemental Figure 2. Night Sweats Decrease Over Time in Both Sexes with A Steeper Slope of Decline and Higher Baseline in Women.**

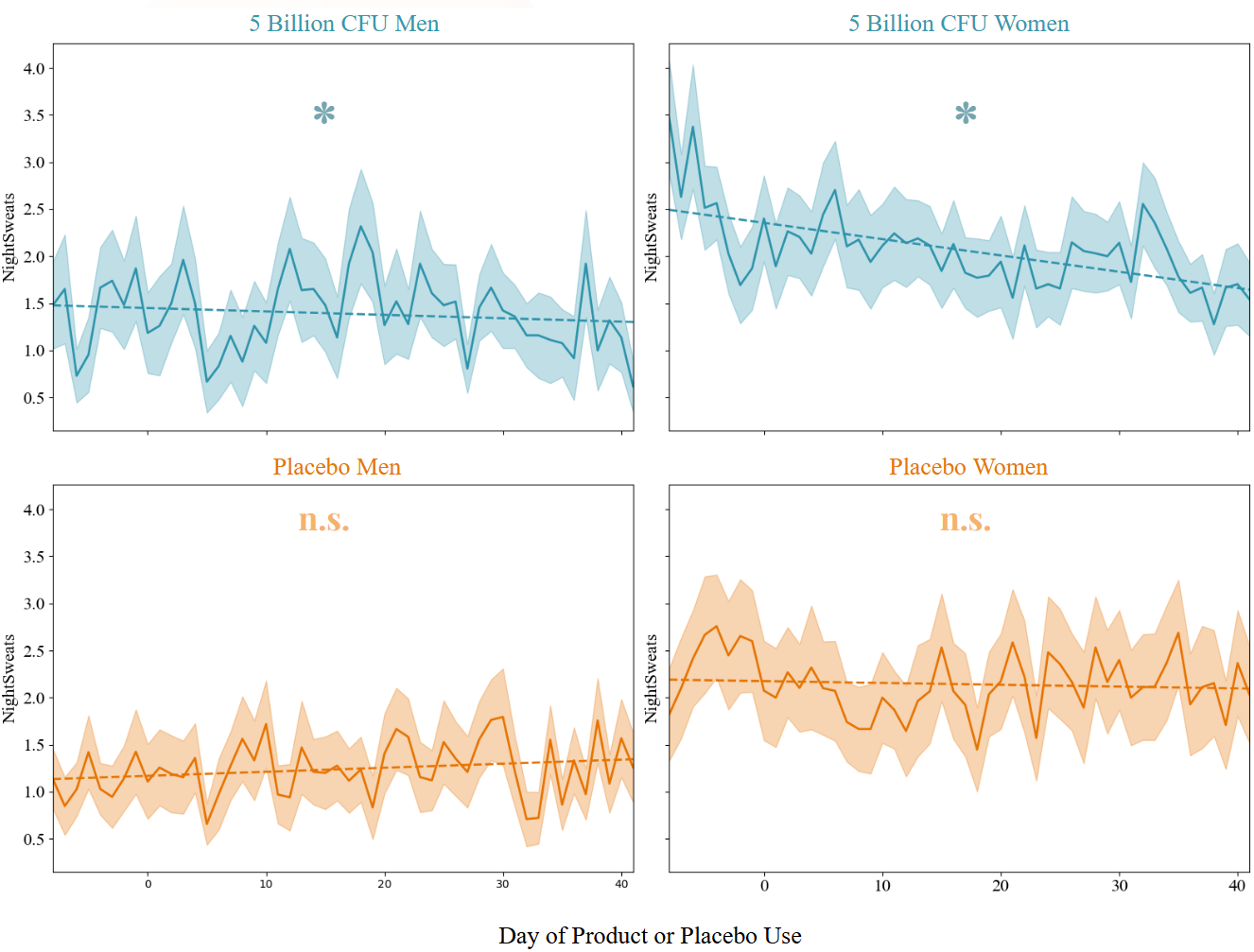

**Supplemental Figure 2. Night Sweats Decrease Significantly in 5 Billion CFU.** Mean ± SEM of daily ratings of night sweat severity (0-5) in 5 Billion CFU (top, blue) and placebo (bottom, orange). Stars indicate statistically significant Mann-Kendall trend over time (p<0.05).

**Supplemental Figure 3. Oura Ring Sleep Metrics Reveal Emergent Group Differences in Sleep Duration Metrics and Baseline Group Differences in Breathing Rate, Heart Rate and Heart Rate Variability.**

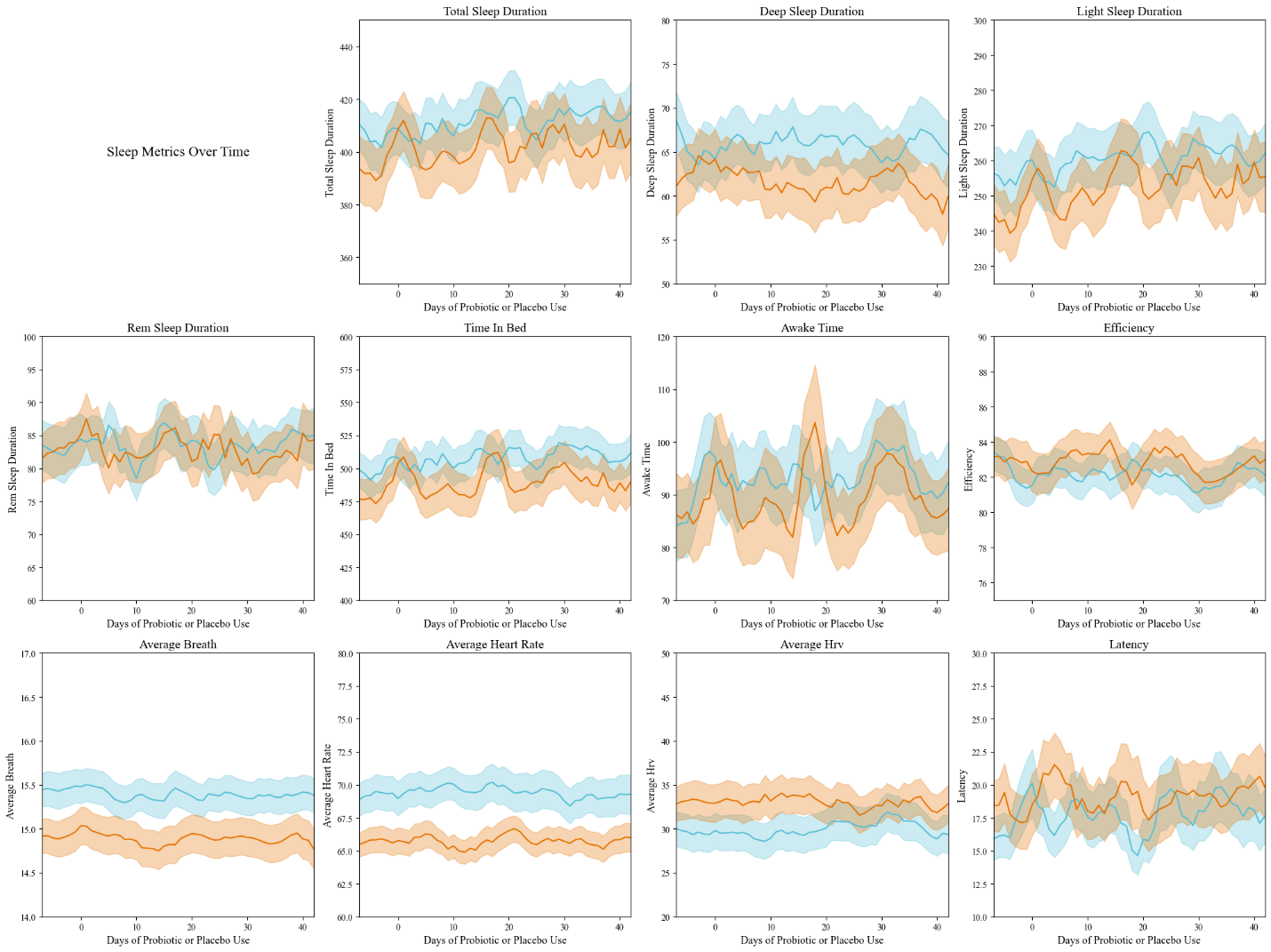

**Supplemental Figure 3. Oura Sleep Metrics.** Mean ± SEM of total sleep duration, deep sleep duration, light sleep duration, REM sleep duration, time in bed, time awake during sleep, sleep efficiency, average nightly breathing rate, average nightly heart rate, average nightly heart rate variability and sleep latency in 5 Billion CFU (blue) and Placebo (orange).

**Supplemental Figure 4. Individual Participants’ Reference-Range Normalized Urinary GABA Concentrations**

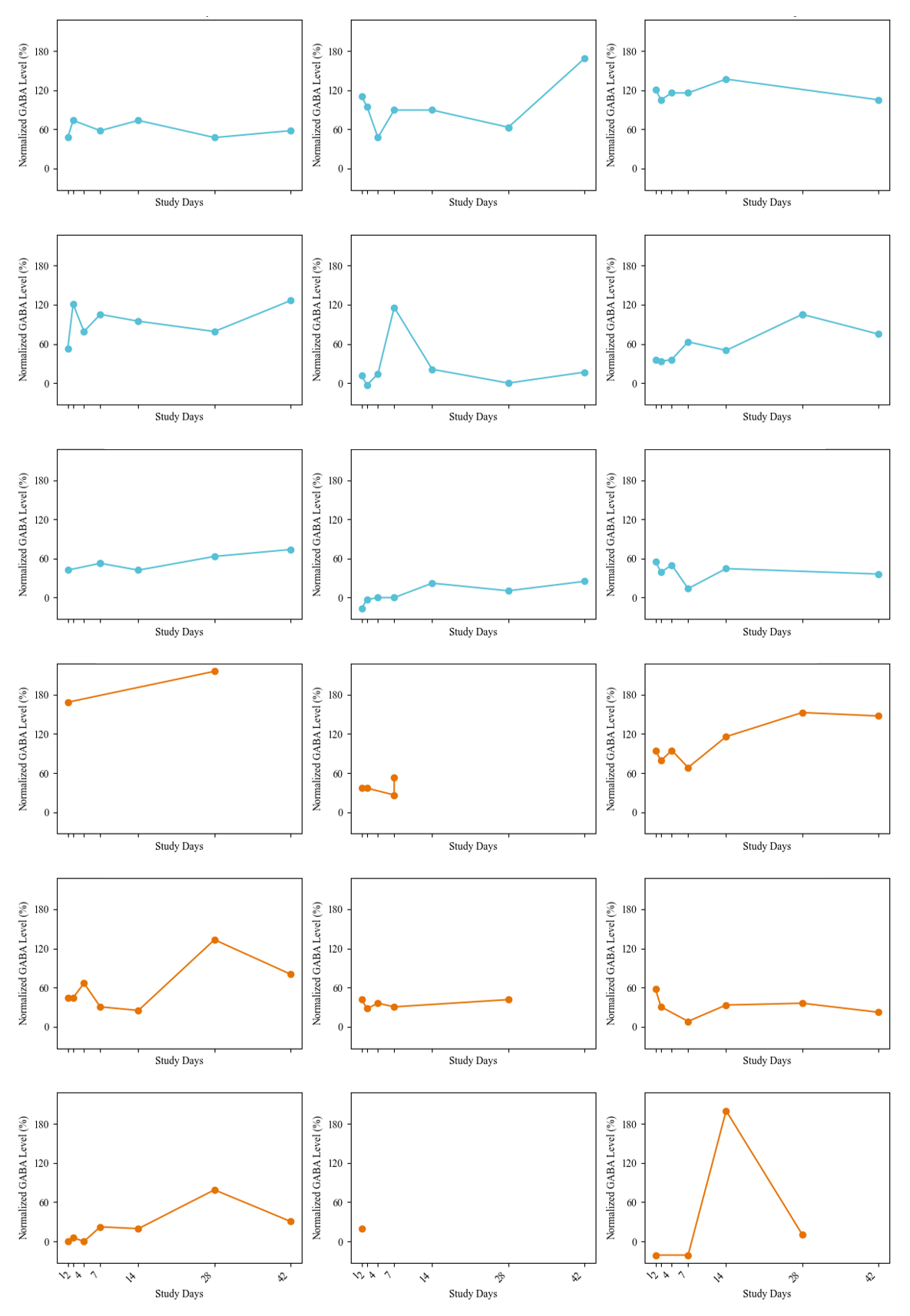

**Supplemental Figure 4. Individual Participants’ Reference-Range Normalized Urinary GABA Concentrations.** Missing days are linearly interpolated. 5 Billion CFU participants are shown in blue and placebo participants in orange. Participants were omitted from group level analyses in the main text if their baseline GABA level was above the normal reference range. Note that several samples in the placebo group were not available for analysis due to spillage during return shipment.

**Supplemental Figure 5. Insomnia Severity Index in Sub Study Participants.**

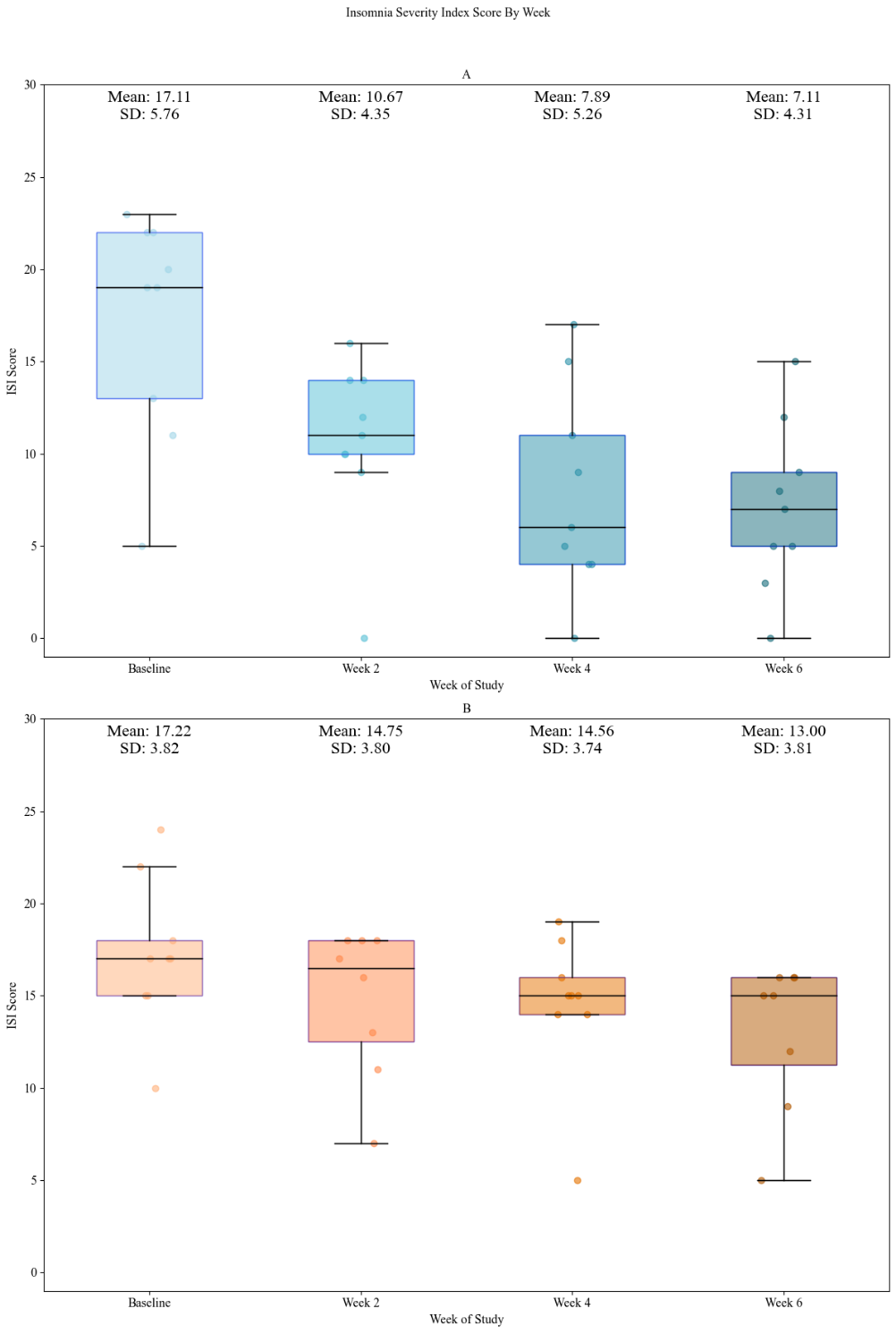

**Supplemental Figure 5. Insomnia Severity Index in Sub Study Participants.** Box plots are overlaid with dots representing individuals’ ISI scores at baseline, Week 2, Week 4 and Week 6. Means and SDs above upper whiskers represent change for that group at that time point. 5 Billion CFU is depicted in blue (top) and placebo in orange (bottom).

**Supplemental Figure 6. GAD-7 Anxiety Score in Sub Study Participants.**

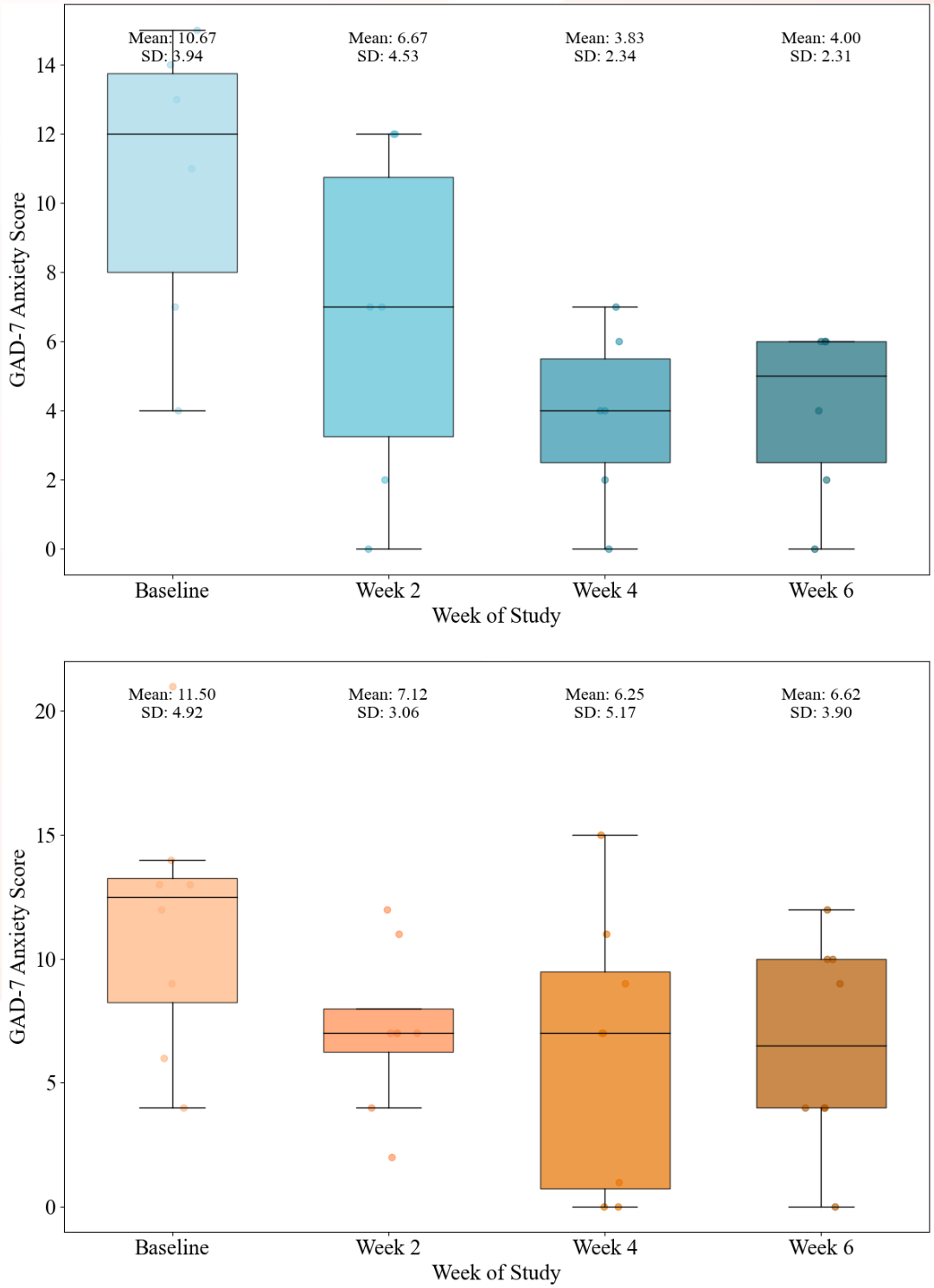

**Supplemental Figure 6. GAD-7 Score in Sub Study Participants.** Box plots are overlaid with dots representing individuals’ GAD-7 scores at baseline, Week 2, Week 4 and Week 6. Means and SDs above upper whiskers represent change for that group at that time point. 5 Billion CFU is depicted in blue (top) and placebo in orange (bottom).

**Supplemental Figure 7. Trajectory of Night Sweat Severity in Sub Study Participants**

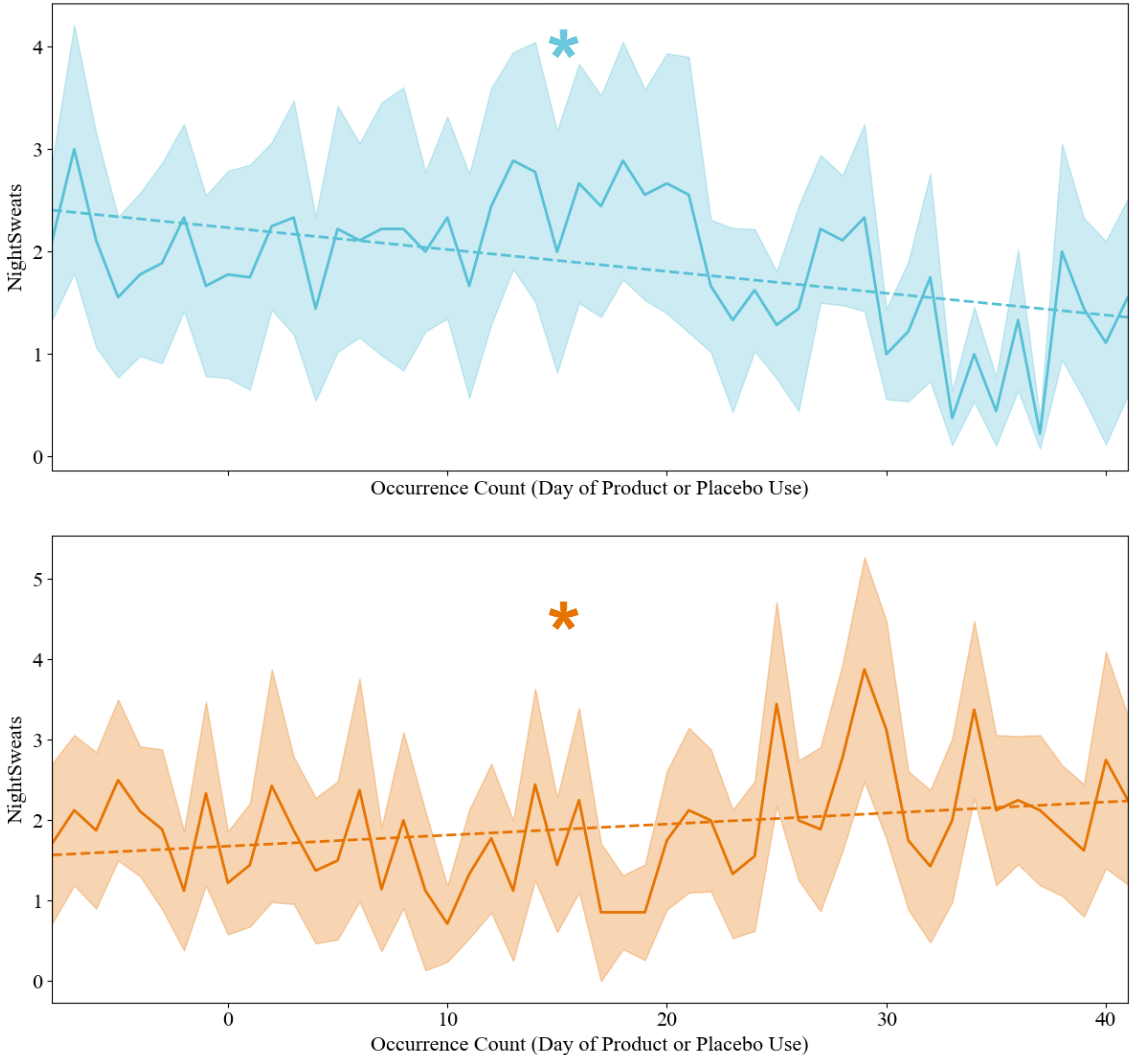

**Supplemental Figure 7. Trajectory of Night Sweat Severity in Sub Study Participants.** Mean ± SEM of daily ratings of night sweat severity (0-5) in 5 Billion CFU (top, blue) and placebo (bottom, orange). 0 Indicated no night sweats, 5 indicated the most severe night sweats. Stars and trend lines indicated statistically significant Mann-Kendall trends over time, decreasing in 5 Billion CFU and increasing in placebo.

**Supplemental Figure 8. Comparison of Reference-Range Normalized Neurotransmitter Concentrations Across the First Week of 5 Billion CFU Lp815 or Placebo Use**

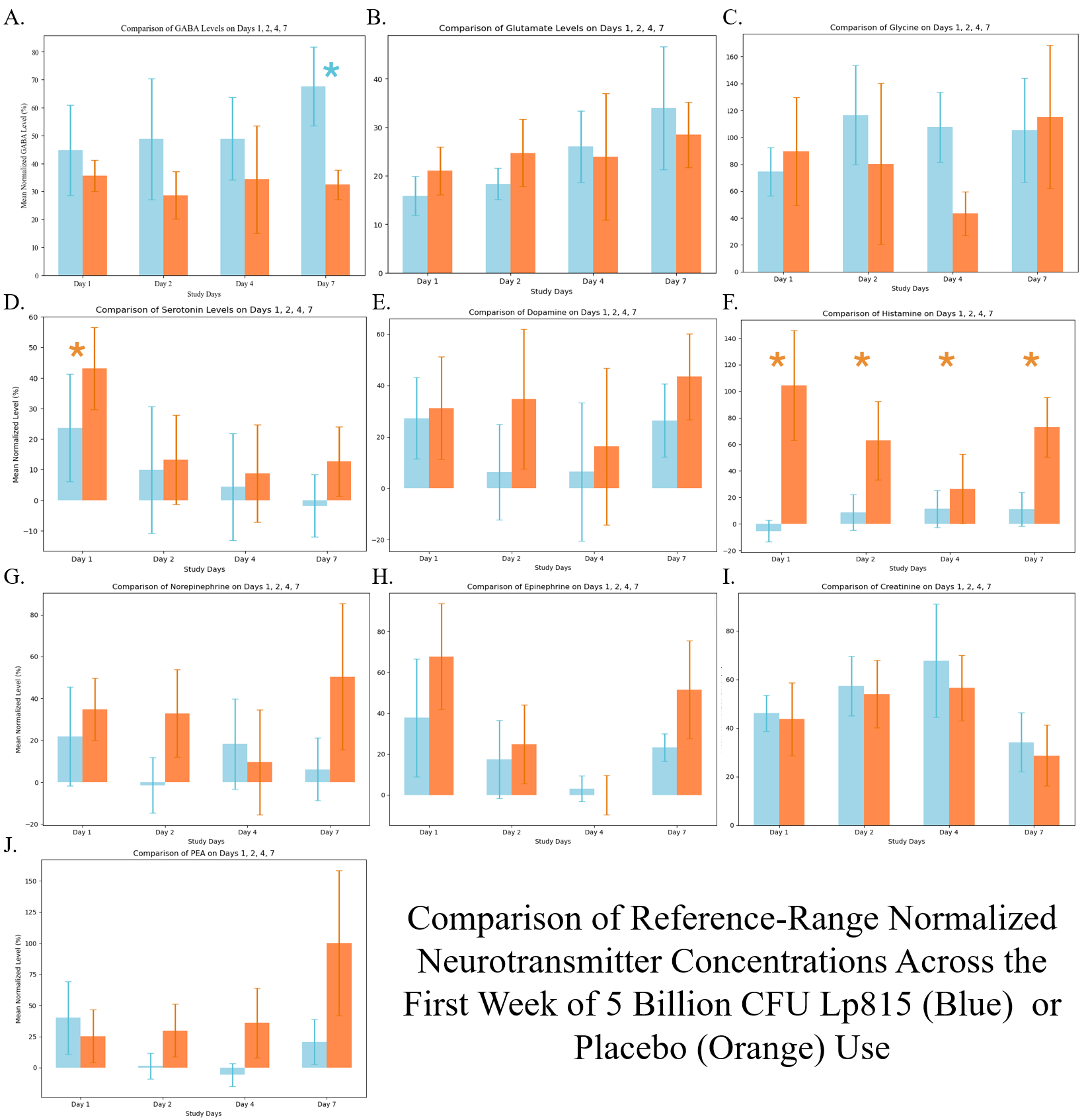

Supplemental Figure 8. Reference-Range Normalized Neurotransmitter Concentrations Across First week. Mean ± SEM of each neurotransmitter’s percent. of reference range. GABA (A), Glutamate (B), Glycine (C), Serotonin (D), Dopamine (E ), Histamine (F), Norepinephrine (G), Epinephrine (H), Creatinine (I), and PEA (J). 5 Billion CFU is shown in blue and placebo in orange. Stars indicate statistically significant difference between groups on the day shown.
